# Artificial Intelligence for Short-Term Modified Rankin Score Prediction after Acute Stroke Symptoms Using Wrist-worn Triaxial Accelerometry Data

**DOI:** 10.1101/2025.04.03.25325214

**Authors:** Benjamin R Kummer, Alexander Gerlach, Shaun Kohli, Joshua Z Willey, Ari Shechter, David S Liebeskind, Girish Nadkarni

## Abstract

**Background:** Functional outcomes after stroke are commonly assessed via modified Rankin Scale (mRS). However, mRS is subject to patient and assessor biases and is impractical to collect in many cases, limiting its impact on post-stroke care. Artificial intelligence (AI) applied to wrist-worn triaxial accelerometry (WWTA) device data can objectively characterize post-stroke functional status and related changes.

**Methods:** We used patient data from REACH Stroke-Sleep, a study investigating WWTA-derived measures of sleep, physical activity, and recurrent stroke risk among patients with acute stroke symptoms. We determined moving accelerometry averages and vector sums over four time windows (minute, hour, day, week). We trained a tree-based (random forest; RF) and deep learning (LSTM) model to predict individual 6-month mRS scores and differences between 1- and 6-month mRS scores. We used 5-fold cross validation, modeled each outcome as binary exact-match between actual-predicted values, and determined area under the receiver-operating curve (AUROC), sensitivity, precision, negative predictive value, and F1 scores for both models. For mRS score differences, we determined mean absolute error (MAE) and standard deviation (SD).

**Results:** We identified 362 patients in REACH Stroke-Sleep, of whom 302 (83.4%) had a 1-month mRS score, 251 (69.3%) had a 6-month mRS score, and 191 (52.8%) had both. Patients wore devices for median 41.0 (IQR 34.4-44.0) days. For all outcomes, RF models (6-month AUROC 0.81, 95%CI 0.74- 0.89; 1-6 month mRS AUROC 0.82, 95%CI 0.76-0.90) outperformed LSTM models (6-month AUROC 0.63, 95%CI 0.55–0.71; 1-6 month mRS AUROC 0.53, 95%CI 0.45-0.61). RF models (MAE 0.37, SD 0.12) outperformed LSTM (MAE 0.87, SD 0.48) for predicting 1-6 month mRS difference, modeled as a non-binarized outcome.

**Conclusions:** We found that AI predicted short-term mRS and mRS changes after acute stroke symptoms from WWTA data with moderate performance. Future studies are warranted to investigate whether multimodal data can improve performance with the goal of developing objective, automatable functional status assessments.

## Introduction

Stroke is a leading cause of long-term disability worldwide.^1,2^ Because most recovery is thought to occur within 3 months after the index stroke event,^3,4^ functional status at 90 days after stroke is a key outcome targeted by many stroke intervention trials and quality improvement efforts. Short-term functional status after stroke is associated with long-term survival, recurrent stroke, post-stroke depression, and healthcare utilization after stroke.^5–13^ While there are several instruments for assessing functional status after stroke, the modified Rankin Scale (mRS) is the most widely used and validated.^14,15,16^

Despite its apparent simplicity and its use as the most widely used tool for functional status assessment after stroke, the mRS is collected in a minority of stroke patients in practice. Indeed, in a large cohort of patients undergoing thrombectomy at a large stroke center in the US, the 90-day mRS was collected in only a quarter of patients ^17^ Difficulties in obtaining mRS assessments are potentially caused by logistical and practical challenges, such as the requirement for contact with trained assessors for completion, as well as language barriers, potentially biased assessor interpretations, and subjective reporting from patients.^18–20^

Another inherent limitation of the mRS is that it is typically applied as a single-point observation.^21,22^ This approach provides a snapshot of disability but fails to capture the temporal evolution of a patient’s functional status over time, thereby providing only limited insights into recovery trajectories and the factors influencing long-term outcomes after stroke. After stroke, patients can experience rapid clinical changes, sometimes portending recurrent stroke, or worsening motor function. Unlike single mRS assessments, dynamic mRS evaluations capture changes over time, enabling interventions to improve quality of life, optimize rehabilitation, adjust medications, and reduce risks of recurrent stroke or rehospitalization. While functional decline is associated with overall mortality and recurrent stroke risk, improvements or decline in mRS may signal actionable events that can impact post-stroke management strategies, therapeutics, and rehabilitative interventions. However, objective data-driven approaches to measure post-stroke functional status change are lacking.

Wearable accelerometry sensors represent a promising avenue for post-stroke functional assessments. Incorporated into smartwatches and smartphones, these devices offer continuous, objective, 3- dimensional measurements of patient mobility and physical activity, both of which are closely related to mRS domains. Moreover, the passive nature of these devices can circumvent the mRS’ requirements for synchronous assessments, as well as subjective barriers.

To our knowledge, no prior attempts have been made to develop predictive mRS models from wearable wrist triaxial accelerometry (WWTA) data. However, due to its ability to process high-granularity, complex data streams with high precision, artificial intelligence (AI) is an excellent approach to accomplish this task. The potential for automating AI models also offers the possibility of further circumventing some of the mRS’ logistical requirements for scheduling synchronous assessments with interviewers. In this study, we aimed to use AI to develop a predictive approach for post-stroke mRS scores at 1 month and 6 months, using WWTA data. Given the ability of mRS score changes to signal meaningful clinical changes in patients after stroke, we also sought to use a similar approach to predict change in short-term mRS between 1 and 6 months after stroke.

## Methods

### Study Design & Patient Population

We used WWTA data collected from a subset of patients in the REACH Stroke-Sleep study. This was a subset of a large prospective study REACH-Stroke (R01HL141494) conducted at Columbia University Vagelos College of Physicians and Surgeons in New York from September 2017 onward.^23^ REACH Stroke-Sleep recruited consecutive adult, English or Spanish-speaking patients who had presented to the emergency department (ED) for stroke or transient ischemic attack (TIA) symptoms. REACH Stroke- Sleep investigates the relationship between patients’ physical activity and sleep in the month after presentation, in addition to subsequent stroke, hospitalization, and mortality over 12 months after presentation. This study excluded patients with a National Institutes of Health Stroke Scale score of 14 or greater, as well as patients whose symptoms precluded completion of protocol, such as current substance abuse, need for immediate psychiatric intervention, or being deemed unable to consent by their ED physician.

In REACH Stroke-Sleep, study patients had the opportunity to opt-in to wear a commercial-grade, FDA 510(k)-exempt multi-sensor accelerometer (GENEActiv ™, ActivInsights Ltd., Cambs, UK) worn on the non-dominant wrist. Patients wore the device for up to 45 days after the enrolling event (with the goal of collecting data over the month following discharge) that captured 6 variables: triaxial accelerometry (x, y, z axes), a vector sum of all 3 axis accelerometry vectors, skin temperature, and light exposure at 10 Hz. All data were compressed into 15-second epochs and recorded in a de-identified manner as a comma-delimited-value list for each patient in a secure web database.

Because we felt that accelerometry was the data stream with the closest relationship with physical activity (and therefore mRS), we only included triaxial accelerometry and vector sum variables in our analysis. As per REACH Stroke-Sleep procedures, clinical and sociodemographic variables were also collected at the time of enrollment, including age, sex, enrolling diagnosis, Charlson Comorbidity Index (CCI), and educational attainment. As part of the REACH Stroke-Sleep research procedures, all participants received a telephone call from a blinded, mRS-trained assessor at 1 and 6 months after the enrolling event to determine respective mRS scores using a simplified version of the mRS (optimized for telephone assessments).^24^ If the patient could not be reached, no mRS score was recorded. Due to data sharing restrictions on identifiable patient information, mRS assessment dates were not available to the study team at Mount Sinai for analysis.

### Measurements

Because most post-stroke recovery is thought to have occurred at 6 months post-stroke, our primary outcome was the mRS score at 6-months after enrollment and we did not utilize the 1-month mRS score as a standalone outcome. Our secondary outcome was the algebraic difference between 1- and 6-month mRS scores (increasing mRS being reflective of functional decline). All outcomes were initially modeled as continuous variables. All procedures were approved by the IRB of Columbia University Irving Medical Center, and as per IRB approvals, de-identified data were shared with the Icahn School of Medicine at Mount Sinai. The IRB of Mount Sinai approved the use of the de-identified patient data shared by Columbia University Irving Medical Center for this research.

### Model Development

We first conducted univariate analyses comparing sociodemographic and clinical variables between 1- month and 6-month mRS score groups, using chi-square and Fisher exact tests for categorical variables and Student’s T-tests or Wilcoxon rank-sum tests for continuous variables where appropriate. Next, we standardized the raw accelerometry data for each axis (“x,” “y,” and “z”) using Z-scoring and applied Gaussian smoothing to reduce noise. We calculated rolling averages of each accelerometry variable as well as the algebraic vector sum of all 3 axes over four-time windows (minute, hour, day, week).

We split the dataset into 80% training and 20% testing sets, which were stratified by mRS scores or score differences depending on the outcome. To address class imbalance, we applied the Synthetic Minority Oversampling Technique (SMOTE) to the training data. We trained two models suitable for granular time-series data analyses to predict all outcomes: a long short-term memory (LSTM) network and a random-forest (RF) model. LSTM is a recurrent neural network architecture, which is a type of deep learning model, whereas RF is a tree-based approach that does not leverage deep learning techniques. We used 5-fold stratified cross-validation for both models, optimizing hyperparameters through grid searches.

### Statistical Analysis and Model Performance Evaluation

Differences between model-predicted and actual values were binarized via the “exact match” method, i.e., model predictions were cast as either equal or different to actual labels, thereby allowing us to determine a 2 x 2 confusion matrix and generate receiver-operating curves (ROC) for each model and both outcomes. Across all folds, we calculated the mean area under the ROC (AUROC) as a metric of overall model performance in classifying each outcome, in addition to mean sensitivity, positive predictive value (PPV), negative predictive values (NPV), and F1 scores for each model. We also performed an additional analysis in which the secondary outcome was modeled strictly as a continuous variable, without exact-matching. In this analysis, we determined the mean absolute error (MAE) between predicted and actual mRS score differences, as well as the MAE standard deviation (SD). All analyses were performed in Python version 3.10. The study data is available by reasonable request to Columbia University Vagelos College of Physicians and Surgeons, with any data sharing being subject to the latter’s institutional board requirements and policies. The Python code for the models in this study can be found at https://github.com/alexgerlach3/mRSWearableAnalysis.

## Results

In the REACH Stroke-sleep cohort, 677 patients consented to GENEActiv monitoring, of whom 524 (77.4%) had analyzable wearable data collected (earliest date September 7^th^, 2017; lates date January 12^th^, 2024). Of these, 362 (69.1%) had at least one mRS score collected, 302 (57.6%) had 1-month mRS scores, 251 (47.9 %) had 6-month mRS scores, and 191 (36.4%) had both 1- and 6-month mRS scores. In the 1-month group, median length of monitoring was 41.0 (IQR 34.4 – 44.0) days, median age was 58 years (IQR 48-68), 167 (55.2%) patients were female, 154 (51.0%) were Hispanic, and 182 (60.3%) were of “other” race. Charlson comorbidity index, enrolling diagnosis, and highest education level were missing to a variable degree in both groups. However, taking these findings into account, neither 1- nor 6-month mRS groups significantly differed in age, sex, ethnicity, enrolling diagnosis, educational attainment, monitoring duration, or mRS score distributions (Table 1).

**Table 1.**
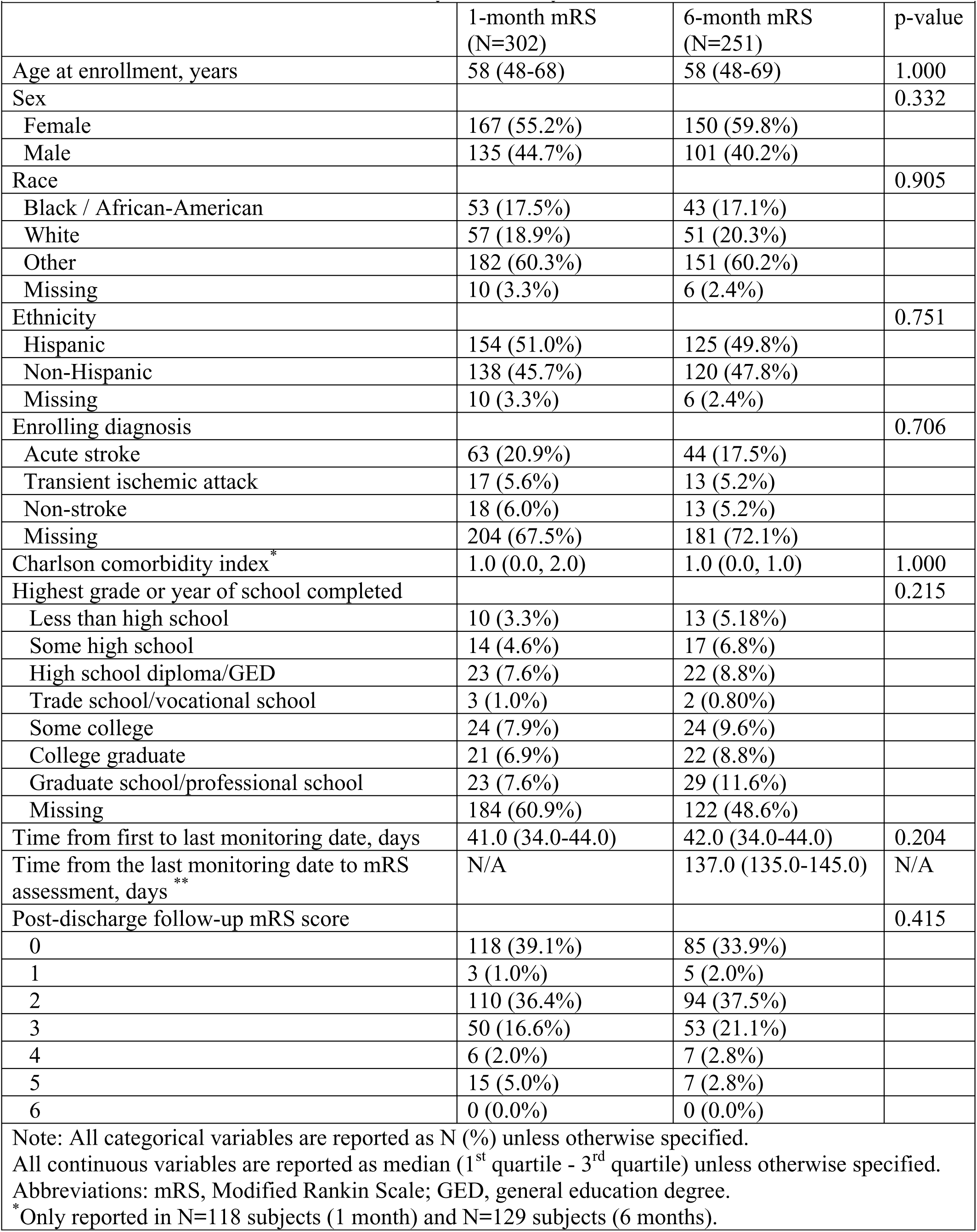
Cohort characteristics, stratified by availability of 1- and 6-month mRS scores.

**Table 2.** Predictive model performance for 6-month mRS scores using triaxial wrist accelerometry.

|  |  |  |  |  |  |
| --- | --- | --- | --- | --- | --- |
|  | **AUROC** | **Sensitivity** | **PPV** | **NPV** | **F1 Score** |
| LSTM | 0.63 (0.55-0.71) | 0.64 (0.45-0.83) | 0.39 (0.35-0.43) | 0.83 (0.78-0.88) | 0.48 (0.42-0.53) |
| RF | 0.81 (0.74-0.89) | 0.65 (0.52-0.77) | 0.42 (0.38-0.46) | 0.85 (0.81-0.89) | 0.50 (0.45-0.55) |
| Note: All metrics were calculated using a binarized “exact-match” approach and reported as mean values across all cross-validation folds with 95% confidence intervals, unless otherwise noted.<br>Abbreviations: AUROC, area under receiver-operating curve; PPV, positive predictive value; NPV, negative predictive value; LSTM, long-short term memory; RF, random forest |  |  |  |  |  |
| **Determined only for 6-month mRS assessments (1-month mRS assessment date occurred prior to the end of monitoring date, resulting in negative time differences). |  |  |  |  |  |

**Table 3.**
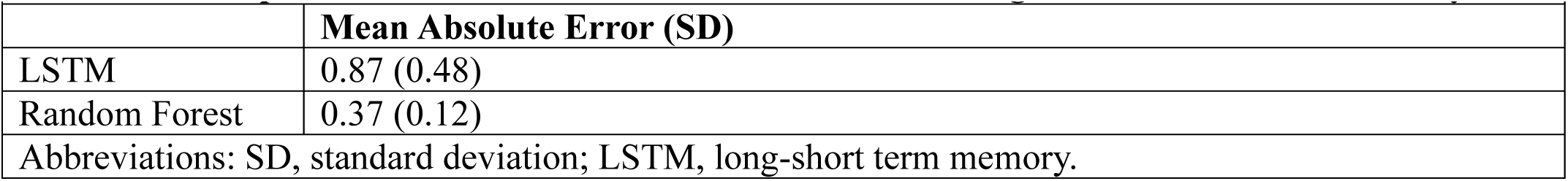
Model performance for 1-6 month mRS difference using triaxial wrist accelerometry.

**Table 4.**
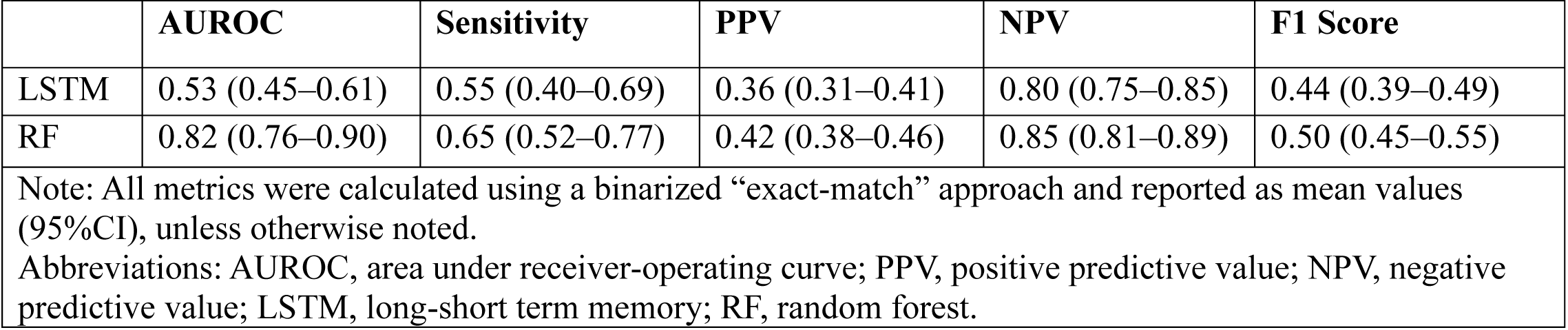
Binarized-output model performance for 1-6 month mRS difference using triaxial wrist accelerometry.

|  |  |  |  |  |  |
| --- | --- | --- | --- | --- | --- |
|  | **AUROC** | **Sensitivity** | **PPV** | **NPV** | **F1 Score** |
| LSTM | 0.53 (0.45–0.61) | 0.55 (0.40–0.69) | 0.36 (0.31–0.41) | 0.80 (0.75–0.85) | 0.44 (0.39–0.49) |
| RF | 0.82 (0.76–0.90) | 0.65 (0.52–0.77) | 0.42 (0.38–0.46) | 0.85 (0.81–0.89) | 0.50 (0.45–0.55) |
| Note: All metrics were calculated using a binarized “exact-match” approach and reported as mean values (95%CI), unless otherwise noted.<br>Abbreviations: AUROC, area under receiver-operating curve; PPV, positive predictive value; NPV, negative predictive value; LSTM, long-short term memory; RF, random forest. |  |  |  |  |  |

In both groups, the most frequently observed proportion of mRS scores was 0 (1-month N=118, 39.1%; 6-month N=85, 33.9%) and 2 (1-month N=110, 36.4%; 6-month N=94, 37.5%) (Table 1). Of the 191 patients with both mRS scores, 36 (18.8%) had numerical decreases in their mRS score (denoting improving functional status), 46 (24.1%) had numerical increases in their mRS scores (denoting worsening functional status), and 109 (57.1%) had no change in their mRS score between these measurements. As a result, the median change in mRS between 1 and 6 months was 0.0 (IQR 0.0-0.0). Among the 46 patients whose functional status worsened, the median mRS increase was 2.0 (IQR 1.0- 2.0) between 1 and 6 months.

Of the two models, the random-forest model demonstrated the higher AUROC for the primary outcome (AUROC 0.81, 95%CI 0.74-0.89) when compared to the LSTM model (6-month AUROC 0.63, 95%CI 0.55-0.71). When modeling the 1-6 month mRS difference outcome as a binary variable via exact-match approach, the random forest model (AUROC 0.82, 95%CI 0.76-0.90) demonstrated superior performance to the LSTM model (AUROC 0.53, 95%CI 0.45-0.61). When modeling the 1-6 month mRS score difference outcome without exact-matching, random-forest models (MAE 0.37, SD 0.12) again outperformed LSTM models (MAE 0.87, SD 0.48) (Figures 1-4).

**Figure 1.**
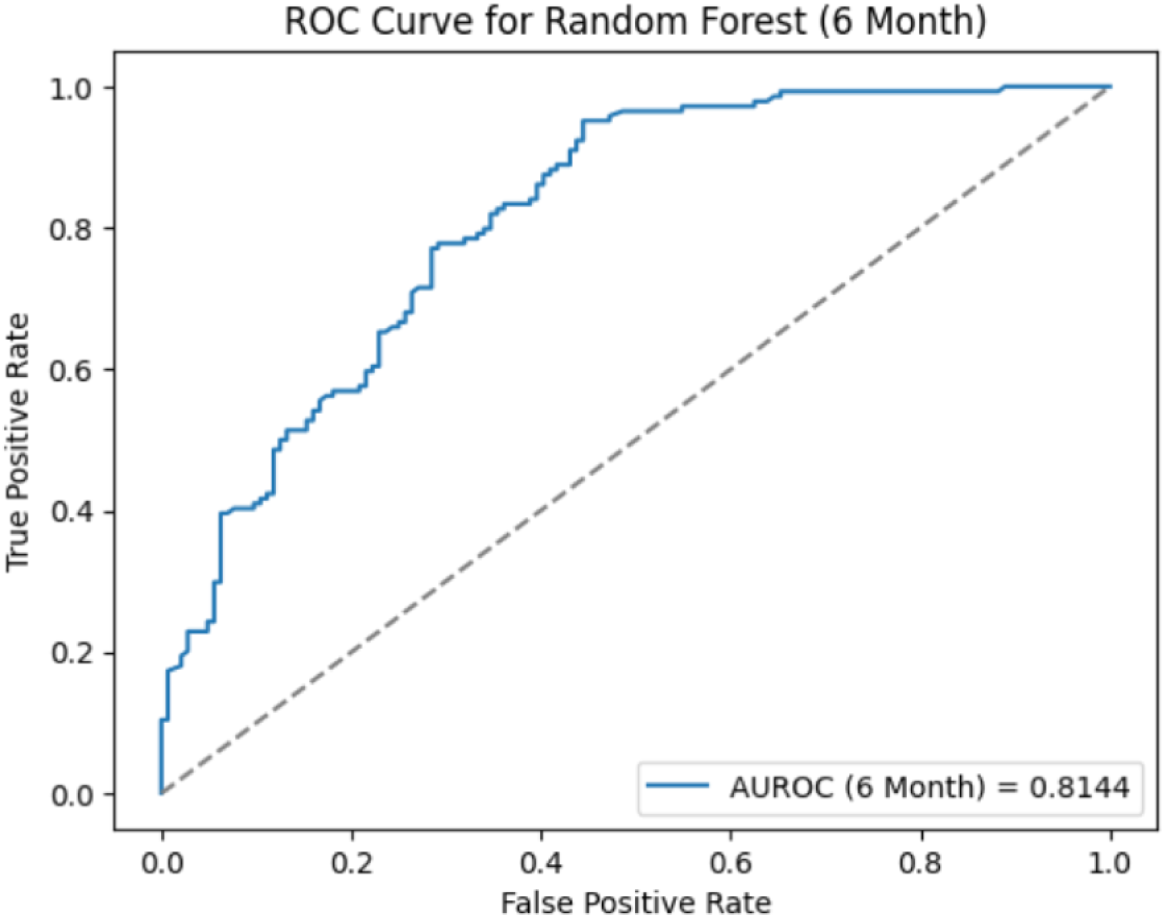
Random forest receiver-operating curve for primary outcome prediction (binarized).

**Figure 2.**
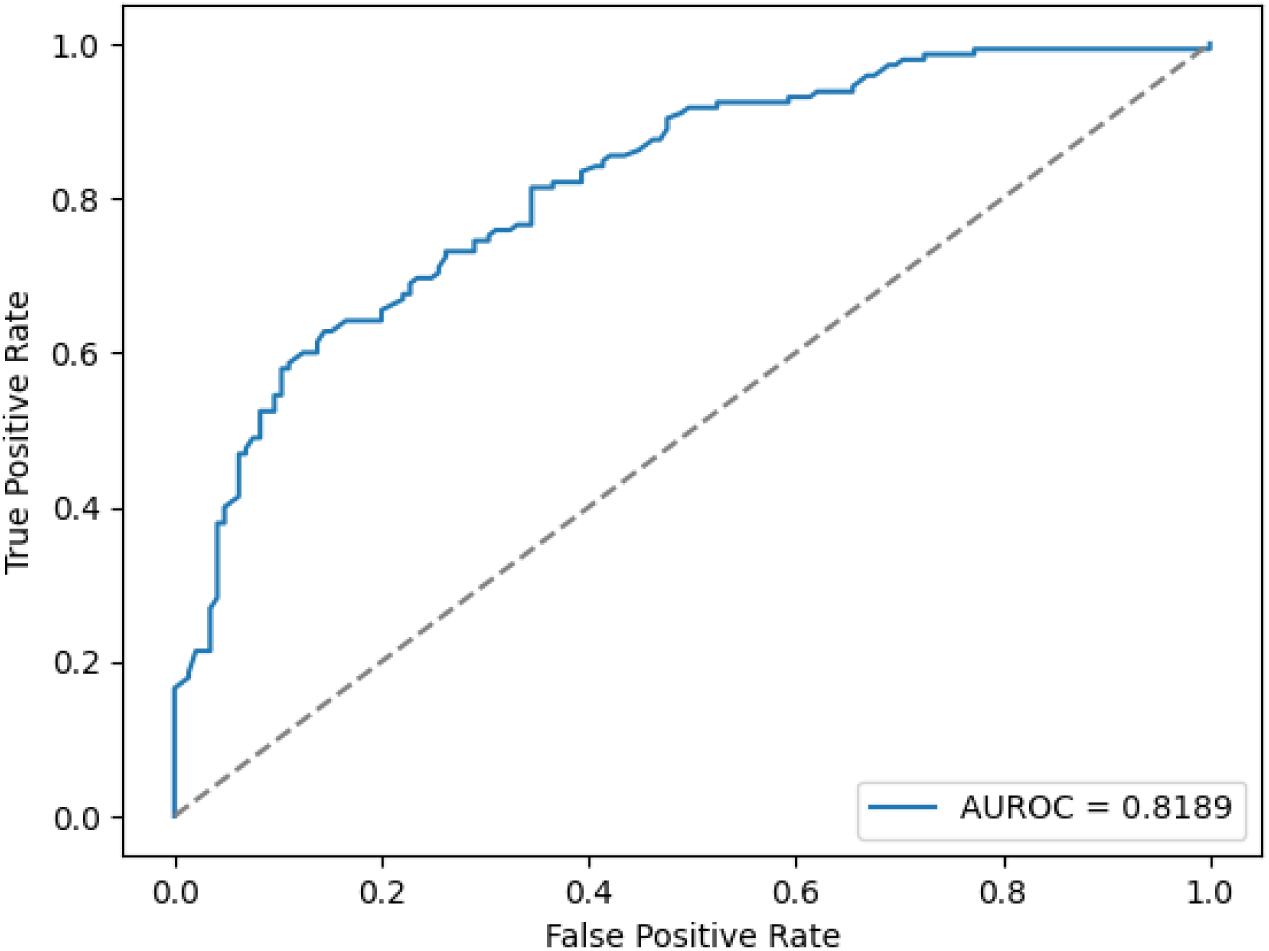
Random forest receiver-operating curve for 1-6 month mRS difference prediction (binarized).

**Figure 3.**
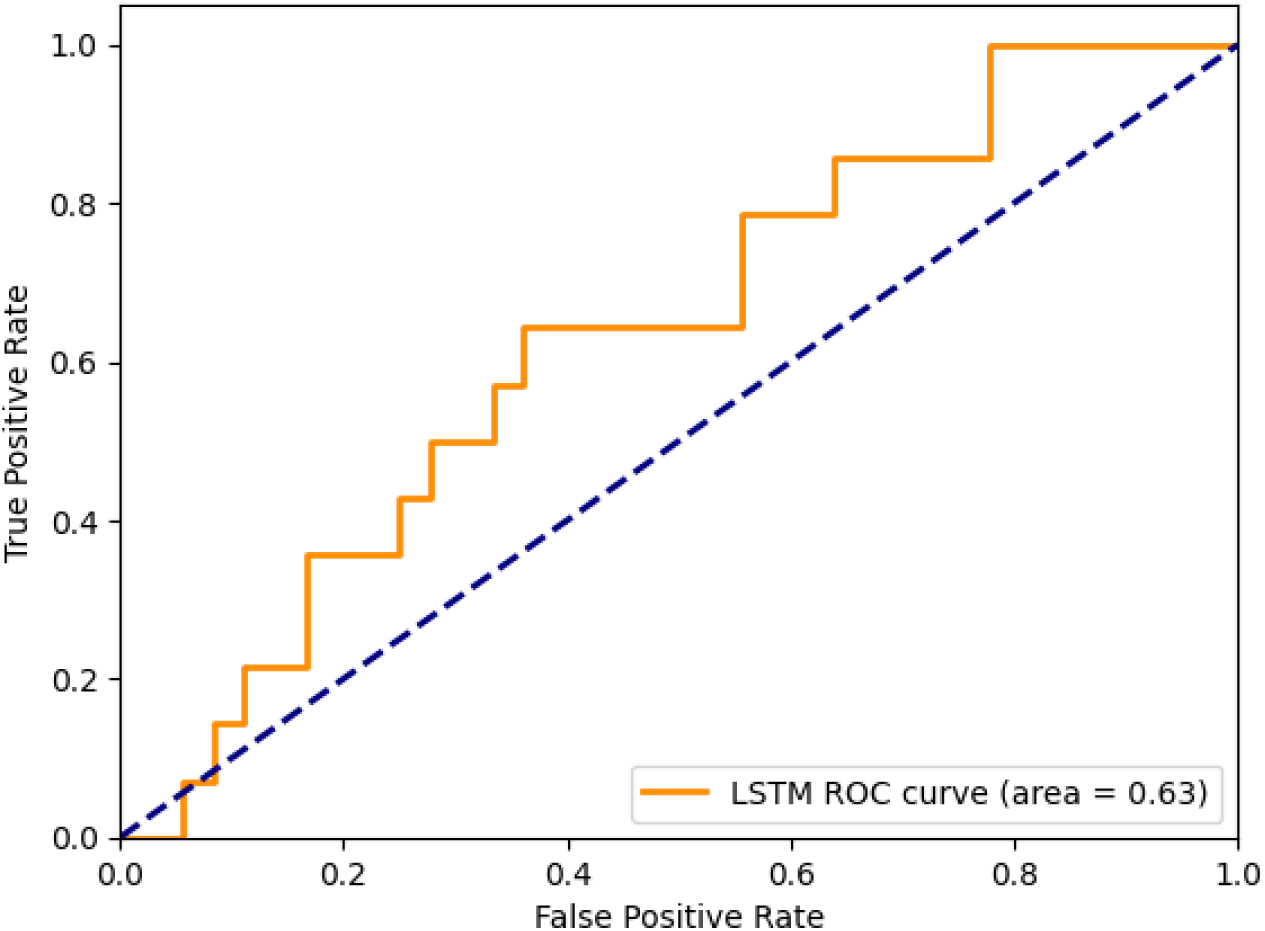
LSTM receiver-operating curve for primary outcome prediction (binarized).

**Figure 4.**
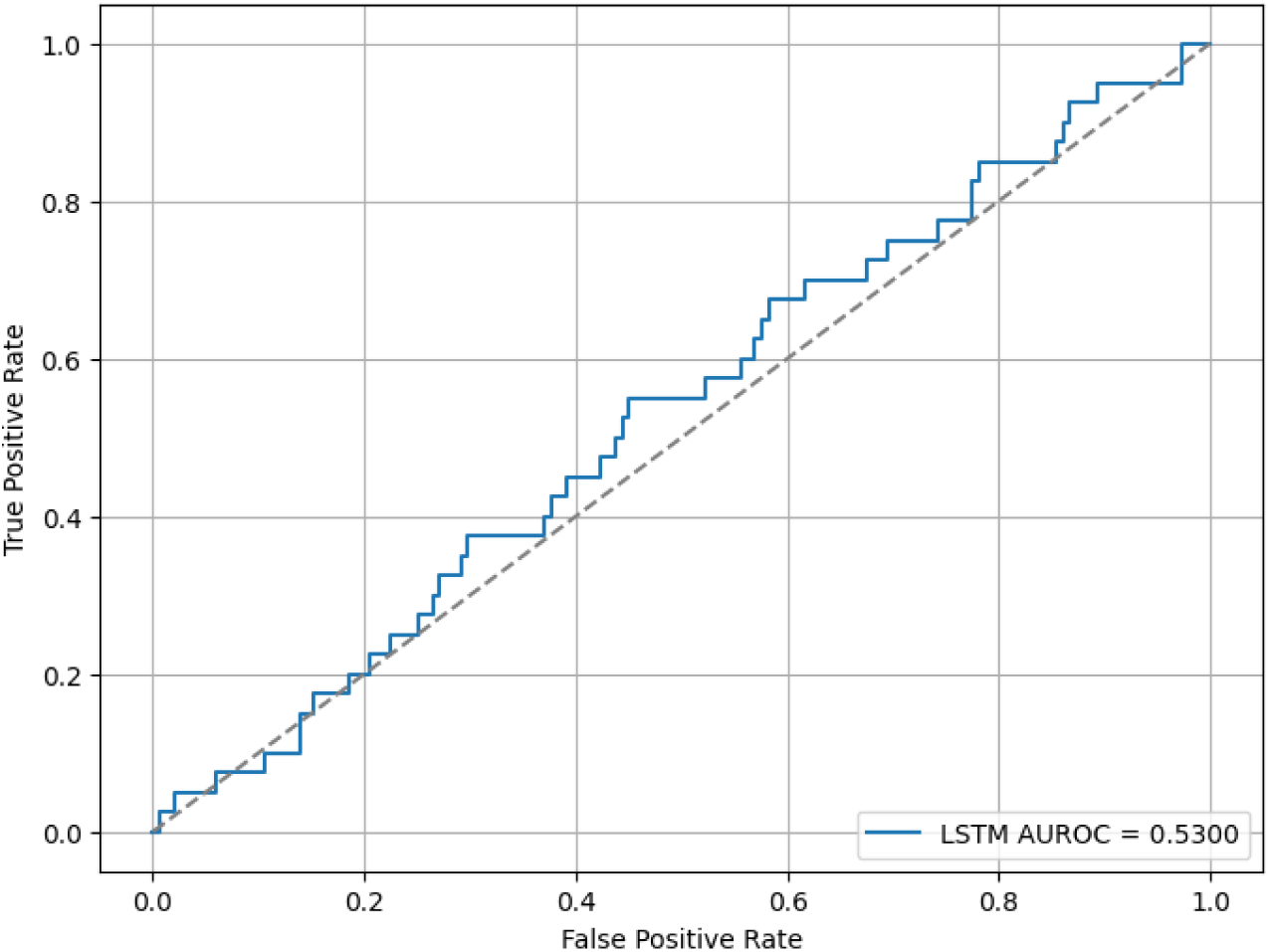
LSTM receiver-operating curve for 1-6 month mRS difference prediction (binarized).

## Discussion

In this retrospective analysis of nearly 300 patients who presented to the hospital with suspected symptoms of acute stroke, we found that two separate ML models could predict short-term functional outcomes as measured by the mRS score at 6 months from WWTA data with moderate-to-good performance, as well as functional status change between 1 and 6 months. The RF classifier, a more readily explainable model architecture than deep learning models, demonstrated a greater overall predictive performance than LSTM for both outcomes across all metrics. To our knowledge, this is the first study to leverage continuous accelerometry data from commercial-grade wearable devices in a post-stroke population to predict short-term mRS and mRS changes after stroke, which can be associated with actionable clinical insights. These findings support the potential of AI in predicting post-stroke functional status applied to WWTA data.

Our findings underscore several advantages of using wearable technology for post-stroke monitoring. First, WWTA data is a passive and continuous measure that reduces dependency on patient self-reporting and clinician-administered assessments, which are often affected by recall bias, subjective interpretation, and barriers including cultural and language differences. Second, accelerometry-based mRS predictions allow for near-real-time monitoring of functional status, potentially enabling earlier interventions for patients whose recovery trajectories show worsening in functional status. Third, the time-resolved accelerometry metrics captured by our model reflect activities of daily living and physical activity. ^25,26^ While physical activity is traditionally most closely assessed and monitored in the post-stroke setting by physical or occupational therapists (PT/OT), this information capture generally occurs during the transition period from hospital discharge prior to visits with stroke program coordinators, stroke providers or primary care providers. Our solution could potentially “close the loop” between discharging and follow-up stroke providers and monitoring PT/OT services to gain better insights into clinical progress after stroke.

Fourth, mRS functional status assessments require either face-to-face contact (in traditional mRS assessment) or synchronous telephone conversation (in the case of simplified mRS) with trained clinicians or coordinators. WWTA-based mRS scores can circumvent this requirement, which can pose significant practical challenges to socially disadvantaged patient populations that may have limited access to telephony, transportation, and/or cellular access. Finally, given that the mRS is tied closely to physical activity, WWTA data can not only surmise a patient’s overall functional recovery but may also serve as a surrogate marker for cardiovascular risk, ^27^ or risk for diseases that disproportionately affect patients who have suffered a stroke, such as deep venous thrombosis or pneumonia.^28^ Our results suggest that non-linear, non-parametric tree-based models may be better suited to effectively capture meaningful clinical patterns within high-dimensional wearable data when compared to recurrent neural networks such as LSTMs.

These results also open promising areas of future research. Beyond physical activity and WWTA data, wearable devices frequently capture additional physiological data including heart rate, blood oxygenation, respiratory rate, sleep duration and quality, light exposure, and skin electrodermal activity, among others. These variables can quantitate cardiovascular reserve^29^ and autonomic nervous system function,^30^ both of which are tied to activities of daily living and can provide additional insight into a stroke patient’s functional status after stroke. Our RF model’s moderate performance in predicting mRS outcomes raises the possibility that incorporating additional sensor data in a “multimodal” approach may improve the predictive performance of our RF model.

Our study is not without limitations. First, many covariables were missing in the overall GENEActiv cohort, including the enrolling diagnosis (which included stroke mimics). However, this missingness is likely random given that there were no programmatic reasons for administering the wearable device to specific population subsets and therefore was unlikely to have biased the results. Furthermore, it is important to note that these patient-level variables not included in our prediction modelling, although future studies should consider integrating multimodal data inputs, including sociodemographic and clinical characteristics, which could refine and potentially improve model performance.

Second, this study’s reliance on a retrospective dataset introduces the potential for unmeasured confounders that may impact mRS prediction accuracy. However, preliminary analysis of the WWTA data streams did not detect any period of missing wearable data (data not shown). Finally, the data set demonstrated notable class imbalance, with over 90% of the cohort having a “favorable” mRS under both 0-3 and 0-2 dichotomization definitions. Although we used SMOTE to handle class imbalances, this may have introduced some bias in overfitting the model, thereby slightly limiting the applicability of our findings.

In conclusion, AI leveraging data from wrist-worn accelerometry devices offers a promising, scalable solution for predicting mRS-based functional outcomes after ischemic stroke. With further validation in prospective cohorts and addition of further multi-modal data, such approaches could be instrumental in augmenting traditional stroke follow-up protocols, ultimately facilitating personalized rehabilitation strategies and optimizing post-stroke care.

## Data Availability

The data used for study is available after reasonable request to Columbia University Vagelos College of Physicians and Surgeons and is subject to the latter?s institutional board requirements and policies.

## Acknowledgments

The authors would like to thank Dr. Ilana Lefkovitz for her help with conceptualizing the analysis of the wearable data, Dr. Jeffrey Birk for his help with sharing the study data, and Dr. Talea Cornelius for her help in providing the cohort variables.

